# Testing the phenotypic decanalization hypothesis: social determinants of hyperglycemia and type 2 diabetes in adult urban Argentinian population

**DOI:** 10.1101/2021.12.24.21268333

**Authors:** María Alejandra Petino Zappala, Guillermo Folguera, Santiago Benitez-Vieyra

## Abstract

Type 2 diabetes, one of the major causes of death and disability worldwide, is characterized by problems in the homeostasis of blood glucose. Current preventive policies focus mainly on individual behaviors (diet, exercise, salt and alcohol consumption). Recent hypotheses state that the higher incidence of metabolic disease in some human populations may be related to phenotypic decanalization causing a heightened phenotypic variance in response to unusual or stressful environmental conditions, although the nature of these conditions is under debate. Our aim was to explore variability patterns of fasting blood glucose to test phenotypic decanalization as a possible explanation of heightened prevalence for type 2 diabetes in some groups and to detect variables associated with its variance using a nation-wide survey of Argentinian adult population. We found patterns of higher local variance for fasting glycemia associated with lower income and educational attainment. We detected no meaningful association of glycemia or its variability with covariates related to individual behaviors (diet, physical activity, salt or alcohol consumption). Our results were consistent with the decanalization hypothesis for fasting glycemia, which appears associated to socioeconomic disadvantage. We therefore propose changes in public policy and discuss the implications for data gathering and further analyses.

## Introduction

Diabetes is a health condition of global concern that afflicts an estimate of 422 million people worldwide, mostly from low-and middle-income countries [1], and is predicted to become a problem affecting one-third of the world population in the next generations [2]. This chronic metabolic condition is characterized by high levels of blood sugar, which can cause heart disease, nerve damage, kidney failure, vision loss, problems during pregnancy, tissue damage requiring leg amputation, and generally increase the risk of disease complications and premature death [1, 3]. In addition, in the last year, a higher susceptibility to adverse outcomes for COVID-19 has been reported for people with diabetes or high blood sugar [4, 5].

Type 2 diabetes, the most common kind [6], is caused by insulin resistance or a lack of production of this hormone. Patients usually require medication, changes in the diet, physical activity and regular checking to maintain their blood sugar levels on a healthy range.

Much work has been done to uncover and describe risk factors for type 2 diabetes or high blood sugar and most public health approaches focus on the spread of information about “healthy habits”, i.e. adequate consumption of fruits and vegetables, regular physical activity, avoidance of tobacco, alcohol and salty foods, and weight control [1, 3]. Although socioeconomic factors related to increased prevalence of diabetes have been described, it is assumed that their effect occurs through the lack of resources (income, time and/or information) to exercise the aforementioned recommendations [3].

### Decanalization

One of the hypotheses seeking to explain complex diseases such as type 2 diabetes points to the influence of environmental stressors in phenotypic variance. This hypothesis is based on the phenomenon of canalization, a concept first coined by Waddington [7] to explain the adoption of discrete and fixed “fates” by cell populations; it was later extended to groups of individuals, to describe their relative phenotypic uniformity despite genetic or minor environmental changes (in this sense, canalization is said to comprise both genetic and environmental mechanisms [8]). In populations living in relatively stable environments, it is expected that stabilizing selection leads to the accumulation of epistatic interactions that restrict the population’s phenotypic variance for adaptive traits near the optima for the environmental range [8]. This reduction of variance means the trait would become “canalized” around these phenotypic values, and as a consequence a majority of the individuals within the population would present similar phenotypes spanning a restricted range. However, under stressful or uncommon environmental conditions, those epistatic interactions may become perturbed; thus, more individuals are expected present extreme phenotypic values, leading to an augmentation of phenotypic variance at the level of the population (“decanalization”), a principle first described by Schmalhausen [9]. Here we will focus on this particular meaning of canalization, a term usually designing a population feature which stems from the robustness of individuals’ developments in the face of environmental change.

Figure 1 summarizes the expectations under the decanalization hypothesis for complex diseases. In stable environments, important physiological variables are tightly regulated in most individuals, so that most of them present trait values near the phenotypic optimum; i.e. the phenotype is canalized, and few individuals present extreme (possibly pathological) values. In populations subject to unusual or stressful environments, phenotypic decanalization would lead, even possibly in the absence of a change in the mean value for the trait, to a higher proportion of the individuals presenting extreme phenotypic values, with possible consequences for their health and life quality.

**Fig. 1.**
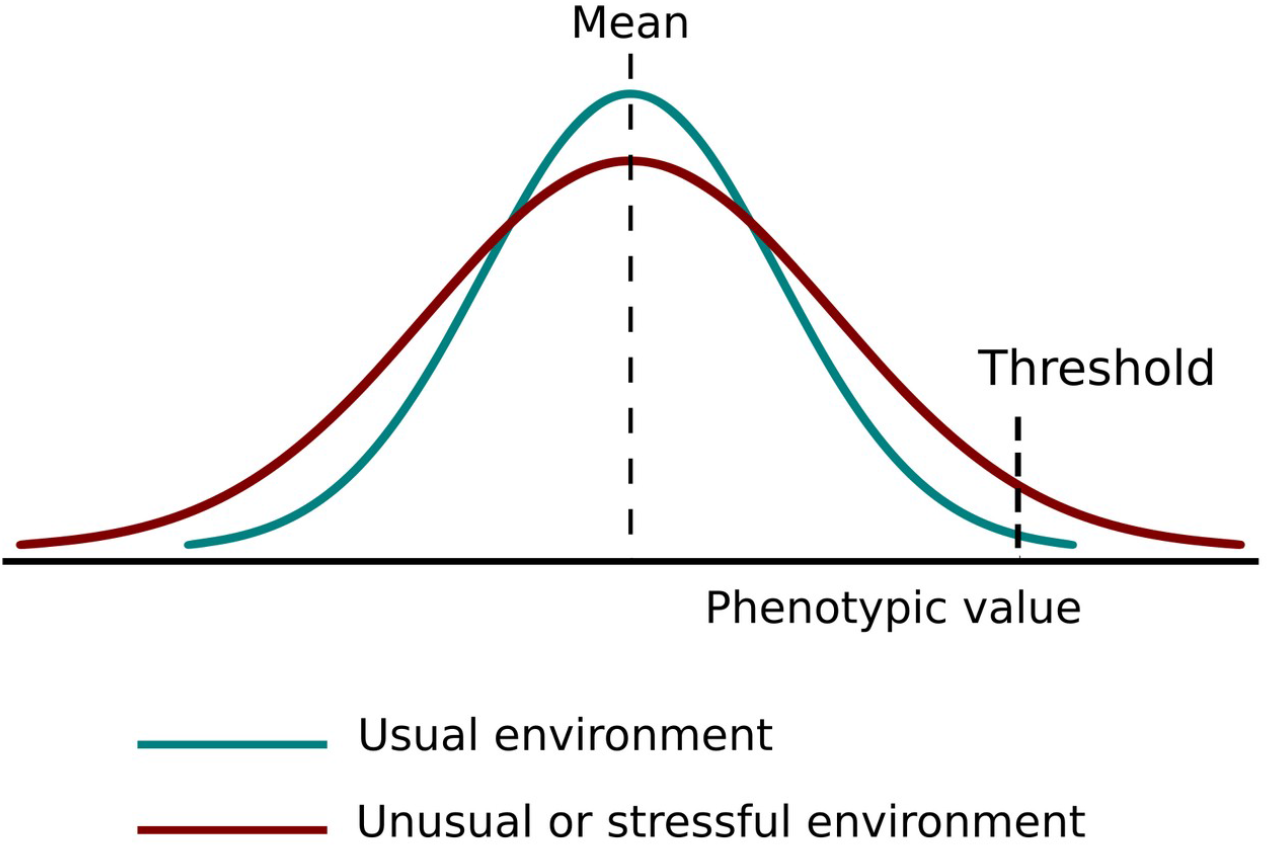
Representation of phenotypic (de)canalization. Expected distribution for the trait values in a hypothetical population under usual and unusual or stressful environments, determining canalized and decanalized phenotypic distributions. A heightened phenotypic variance results in a higher proportion of individuals with extreme phenotypic values at both ends of the distribution.

There are two different accounts for the decanalization hypothesis that can be applied to metabolic diseases such as diabetes in human populations. Both would consider the disease prevalence, in this case type 2 diabetes, as related to the phenotypic decanalization of an otherwise canalized trait (here, the homeostatic mechanism regulating blood glucose levels) in some groups within the population. They differ, however, in terms of which are the relevant environmental factors causing this decanalization. One of the hypotheses points towards the introduction of industrialized foods and changes in motor activity (i.e. sedentarism) as driving the augmentation of phenotypic variability for blood sugar levels in human populations and, therefore, leading to a higher prevalence of type 2 diabetes [10]. According to this version, it follows that efforts to tackle diabetes should be focused on the promotion of healthy habits [2, 10]. A second version of the decanalization hypothesis was formulated by Lewontin and Levins [11, 12]. The basis of this explanation is similar, but the authors, when discussing human populations, refer to the poor, excluded and marginalized communities suffering from multiple environmental stressors which are not under their control, that depend on the historical relationship of the populations with their environment, including social relations between individuals. According to this view, inequalities directly impact on physiological processes through a multitude of mechanisms (including psychological stress derived from the position within class hierarchies) affecting health as a whole. Under the stressful conditions to which these groups are subjected, mechanisms buffering the phenotype from environmental variation break down, and latent differences between individuals become manifest [11, 13]. According to Himmelstein et al. [13], homeostatic capacity generally erodes with age through the accumulation of stressors, but differences can be found when accounting for factors such as race or income, so that underprivileged people’s homeostatic mechanisms deteriorate faster; therefore these groups present greater phenotypic variability earlier. These authors, and others focusing on the social determinants of health, argue that health policies that rely on the spread of information about healthy habits to tackle diabetes are not the best way to address the problem, and they also shift the responsibility to the individuals when the causes of disease would be of social origin and therefore outside of their control [12, 14]. This version of the hypothesis is not incompatible with other approaches like the “psychosocial stress theory”, although it provides for a possible mechanism through which stimuli of very different kinds which are perceived as stressful (or protective) may affect the susceptibility to suffer from ill health [15].

It is known that physiological levels of fasting blood glucose in humans are tightly regulated; according to current guidelines the healthy range lies between 70 and 100 mg/dL. However, to our knowledge, the decanalization hypothesis for this trait in human populations has not been systematically assessed [16]. Evidence from experimental studies carried out in *Drosophila melanogaster* supports the idea that the likelihood of extreme metabolic phenotypes can be augmented by dietary changes leading to a higher phenotypic variance [17]. There are obvious obstacles to the testing of this hypothesis in humans, although some observational studies show results consistent with it [13, 18].

A corollary to both decanalization hypotheses is that trying to model individual risk for diabetes would be fruitless, as would be looking for a difference in means of blood sugar levels between groups. Therefore, different statistical approaches and, ideally, surveys designed to this end would be needed to identify groups with a higher susceptibility to diabetes and to design public health strategies to counteract this problem, an objective of the utmost importance [3].

### Diabetes in Argentina

In Argentina, according to the last National Surveys on Risk Factors for Noncommunicable Diseases the prevalence of diabetes and hyperglycemia (by self-report) has been on the rise in the last two decades [19]. Public health campaigns at the national and regional level have focused on the promotion of healthy habits [20], and although a heightened prevalence was detected in the surveys for people with lower educational attainment and public (vs. private) health insurance, no mechanism was proposed other than lack of access to or noncompliance with dietary and physical exercise guidelines [19].

In contrast to previous surveys, in order to assess the possible under-diagnosis of diabetes and high blood sugar, the last National Survey carried out in 2018 incorporated biochemical measurements, including capillary fasting blood glucose. In this work, using these determinations, we tested whether data from the Argentinian urban population support the decanalization hypothesis for glycemia, in order to explore the determinants of hyperglycemia and diabetes. Moreover, we were able to explore associations with socioeconomic and individual variables that could account for the differences observed in the patterns of fasting blood glucose. This setting provides us with a basis to discuss current public preventive approaches and propose changes in health policy and future survey design and analysis.

## Materials and methods

### Data

The raw database from the 2018 National Survey on Risk Factors for Noncommunicable Diseases, consisting on several variables from 25,208 adults from all provinces randomly selected through a probabilistic multi-stage sample design, was obtained from the INDEC webpage. All raw data are available at https://www.indec.gob.ar/indec/web/Institucional-Indec-BasesDeDatos-2.

Although information on diabetes diagnosis (by self report) was available, it was not used for these analyses, as it did not allow us to test the decanalization hypothesis. Therefore, we used only the determinations of fasting blood sugar; data were filtered to exclude those cases where these determinations were not performed. The relevant explanatory variables were chosen according to bibliography and availability of data on the survey. Those were age, sex, income, educational attainment for the interviewee and head of the household (grouped in three levels; 1 = elementary school not completed, 2 = completed elementary school, high school not completed, 3 = completed high school), density (ratio of inhabitants / number of rooms on the household), utilities (a joint variable combining the information for water, gas and sewage system availability, rescaled between 0 and 1), paid working time (grouped in three levels, besides zero for unemployed or retired interviewees, and not including unpaid work; 1 = time < 35 weekly hours, 2 = 35 < time < 45 weekly hours, 3 = time > 45 weekly hours), average number of daily fruits and vegetables consumed, levels of alcohol (0 = no problematic consumption, 1 = problematic regular consumption or problematic episodic consumption, 2 = problematic regular consumption and problematic episodic consumption) and salt intake (0 = does not use salt, to 3 = always adds salt to meals), levels of physical activity (1 = low, 2 = intermediate, 3 = high) and sedentarism (daily minutes spent sitting).

### Generation of new variables

We generated a categorical variable for levels of fasting blood sugar (in mg/dL) based on the raw values provided in the survey database. However, instead of the standard ranges of fasting plasma glucose used by convention to diagnose diabetes and prediabetes [1], we implemented a more gradual categorization, with its levels being hypoglycemia (glycemia<70), normal (70<=glycemia<110), borderline (110<=glycemia<140), prediabetes (140<=glycemia<200), diabetes (200<=glycemia<240) and severe diabetes (glycemia>=240).

Age is a well described risk factor for hyperglycemia and type 2 diabetes [19, 21, 22]. To account for its effect on blood sugar levels, we calculated residuals from the linear regression between glycemia and age, and created a variable grouping absolute residuals in three categories: Normal (residual<40), Outlier (40<=residual<80), roughly representing Q1 + IQR and Q3 + IQR, and Extreme (residual>=80), approximately Q1 + 2.5*IQR and Q3 + 2.5*IQR. This variable serves to group individual values that deviate from the estimate in both directions and is therefore useful to detect decanalization.

Monthly household income was also categorized in four levels: high or A (income>=40,000 pesos argentinos), middle or B (20,000<=income<40,000), low or C (10,000<=income<20,000) and very low or D (income<10,000). At the time the survey was performed, the cutoff for indigence for an average family (two adults, two children) in Buenos Aires was of 10,122 pesos, approximately corresponding to 280 US Dollars [23]. In this sense an average family in group D would be considered as indigent.

Finally, Coefficients of Variation for glycemia were calculated as 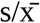, being s and 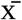 the estimators of standard deviation and mean for each group.

### Statistical analyses

All statistical analyses were performed in Rstudio [24] using R version 3.6.3 [25].

We applied several statistical analyses to test the decanalization and alternative hypotheses. First, we applied a “traditional” approach, i.e. we tested whether average glycemia values per individual can be predicted by the explanatory variables, which were detailed above. Contrarily to the decanalization hypothesis, which would search for an increased within-group variance to detect factors behind increased susceptibility to diabetes, all of these “traditional” analyses are aimed at modeling individual risk for diabetes.

For this approach, we applied supervised machine learning algorithms such as random forest (RF), Gradient Boosting Machine (GBM), and regression Support Vector Machine (SVM). These analyses were performed with randomForest, caret, and e1071 R packages [26-28]. RF works by repeatedly subsampling from the data, constructing a decision tree for each subsample (in the case of regression, the response variable is discretized) and then averaging or combining the resulting trees. GBM also works with trees, by producing an ensemble of “weak” ones into a “strong” classifier, where each decision tree has a different influence according to performance [29]. SVM works by finding a hyperplane defined by the optimal parameters to explain the response variable (i.e. minimizing the distance to the maximum number of points) [30, 31].

Second, to test the predictions of the decanalization hypothesis and to inquire the explanatory variables influencing diabetes susceptibility, we first describe the multivariate space for our data. A Categorical Principal Component Analysis (PCA) was performed using the princals function from the Gifi R package [32], where all explanatory variables excepting sex were considered ordinal. This analysis combines linear multivariate analysis with optimal transformation of the categorical variables using alternating least squares [33]. No variables related to blood sugar level were used as active variables in this analysis. Therefore, the obtained components represented a multivariate space defined by the explanatory variables. Then, on this multivariate space, we mapped the categorical variable that groups absolute residuals from the regression between glycemia and age in three categories: Normal, Outlier, and Extreme, as described above. As we used absolute residuals, those categories represent the variability of glycemia in relation to its expected value. Thus, they are an approximation to groups with different levels of decanalization, each one characterized by broader deviations from expectation. To test whether individuals from the Normal, Outlier, and Extreme groups occupied distinct sectors of the socioeconomic multivariate space, the scores for individuals in all PCA components were subjected to a Permutational Multivariate Analysis of Variance (PERMANOVA [34]) using the adonis function of the vegan R package [35], having previously checked the assumption of multivariate homogeneity of variances (using functions betadisper and permutest from the same package). Bonferroni-adjusted pairwise comparisons were performed with the pairwise.adonis function [36].

In the same multivariate space of socioeconomic explanatory variables, we determined regions of high and low variance for fasting blood glucose. With this aim, we used a procedure similar to a running average in two dimensions. The algorithm determines neighborhoods in the multivariate space and then computes the standard deviation for fasting blood glucose for all the observations within. This function returns a vector containing the running standard deviation for the neighborhood around each observation. This analysis was inspired by spatial analysis: imagine a city where diabetes susceptibility varies among real neighborhoods. Then, following the decanalization hypothesis from Lewontin and Levins [11, 12], marginalized neighborhoods should show higher values of standard deviation in glycemia. Our data sadly lacked detailed geographical information, but through PCA we could construct “neighborhoods” of individuals that share certain socioeconomic characteristics.

After identifying through the PCA and running standard deviation the variables that better explained differences in neighborhood variability for glycemia, we tested whether these variables were associated to different patterns of blood sugar levels. With this aim, we performed a log likelihood ratio test of independence to explore if the individuals with normal, outlier or extreme glucose residual values were equally distributed in the different income and educational attainment groups. using the GTest function from the DescTools package [37].

## Results

First, data from the National Survey were filtered. From a total of 4,477 individuals on which blood glucose was measured, 4,115 individuals were retained for which complete records were available for all variables (Table 1, Additional File 1). Given the reported relationship between glycemia and age, which was also confirmed by our exploratory analyses, we calculated residuals from the linear regression between glycemia and age and also created a categorical variable for groups according to absolute residuals (see Materials and methods).

**Table 1.** Summary for all covariates used as explanatory variables in the analyses

| Covariates | Mean ± SD or % |
| --- | --- |
| Age (years) | 50.2 ± 16.1 |
| Sex | Male: 41.55% |
|  | Female: 58.45% |
| Max. Educational attainment: interviewee | Incomplete elementary school: 8.41% |
|  | Incomplete high school: 33.90% |
|  | Complete high school and above: 57.69% |
| Max. Educational attainment: head of household | Incomplete elementary school: 9.28% |
|  | Incomplete high school: 35.36% |
|  | Complete high school and above: 55.36% |
| Monthly income for the household (Argentine Pesos) | 23,369 ± 18,313 |
| Working time (weekly hours) | No working time: 38.5% |
|  | 0 to 35: 22.2% |
|  | 35 to 45: 23.1% |
|  | > 45: 16.2% |
| Density (inhabitants / household rooms) | 1.00 ± 0.70 |
| Utilities (from 0 = no access to gas, water or sewage system to 1 = access to gas, water and sewage system) | 0.64 ± 0.17 |
| Level of physical activity | Low: 46.2% |
|  | Intermediate: 36.8% |
|  | High: 17.0% |
| Sedentarism (daily minutes spent sitting) | 261.4 ± 174.0 |
| Daily fruit & vegetable consumption (portions) | 2.03 ± 1.62 |
| Salt consumption | Does not add salt to meals: 21.6% |
|  | Sometimes adds salt to meals: 50.8% |
|  | Frequently adds salt to meals: 17.4% |
|  | Always adds salt to meals: 10.2% |
| Alcohol consumption | No problematic consumption: 82.9% |
|  | Regular or episodic problematic consumption: 13.2% |
|  | Regular and episodic problematic consumption: 3.8% |

First, we performed the “traditional” analyses to model raw values or residuals for fasting blood glucose measurements. We found no regression algorithm that could satisfyingly fit our data, as they all explained a low proportion (<10%) of variance for both variables (Additional File 2).

We performed a Categorical Principal Component Analysis and generated a multivariate space of the explanatory individual and socioeconomic variables; we retained 8 components explained 81.1% of the total variance. Loadings for the first 4 components (with a cutoff of 0.1) are shown in Table 2 (for all 8 components retained, see Additional file 3).

**Table 2.**
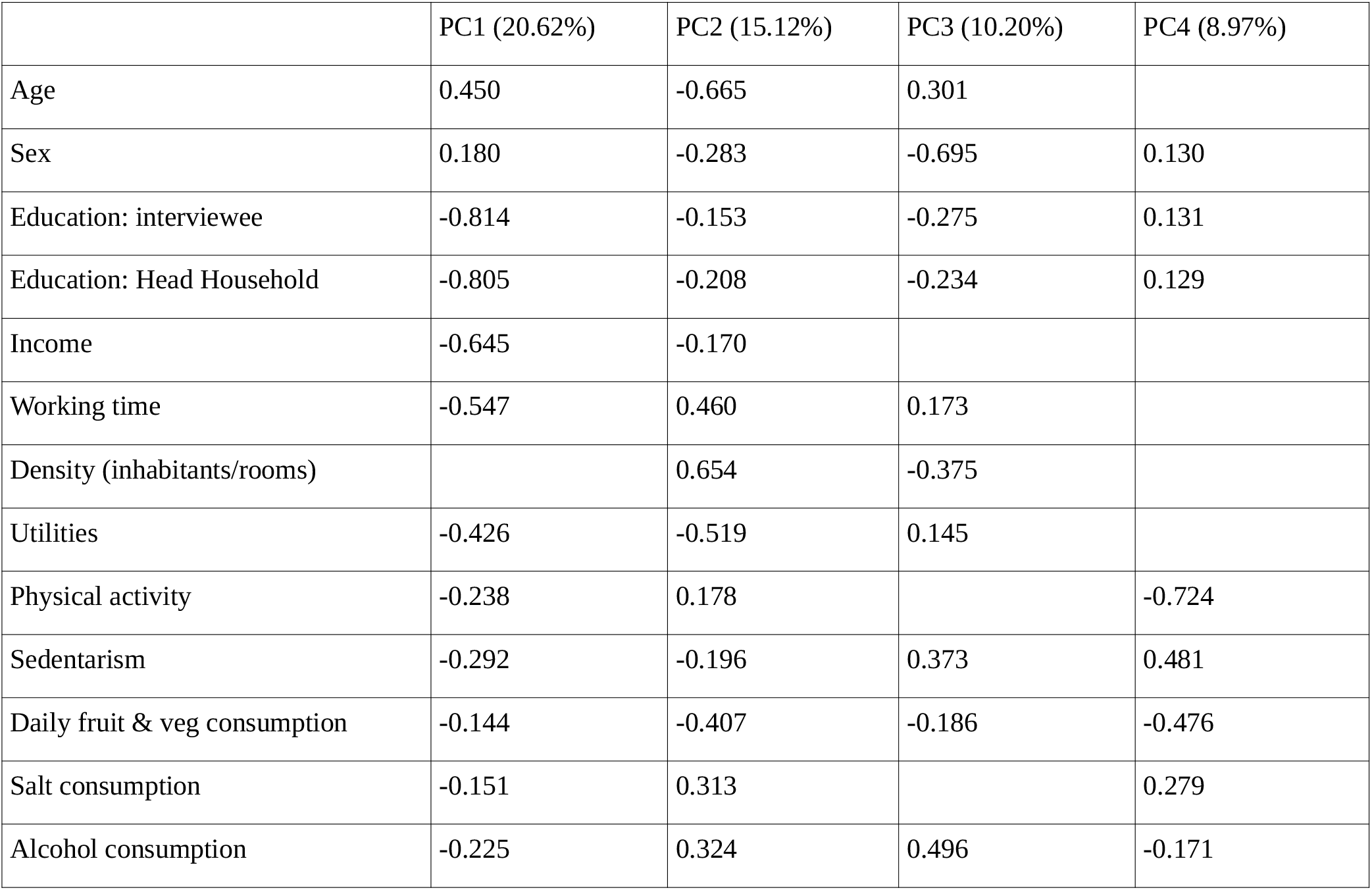
Loadings for all variables in the four Principal Components and % of explained variance.

|  | PC1 (20.62%) | PC2 (15.12%) | PC3 (10.20%) | PC4 (8.97%) |
| --- | --- | --- | --- | --- |
| Age | 0.450 | -0.665 | 0.301 |  |
| Sex | 0.180 | -0.283 | -0.695 | 0.130 |
| Education: interviewee | -0.814 | -0.153 | -0.275 | 0.131 |
| Education: Head Household | -0.805 | -0.208 | -0.234 | 0.129 |
| Income | -0.645 | -0.170 |  |  |
| Working time | -0.547 | 0.460 | 0.173 |  |
| Density (inhabitants/rooms) |  | 0.654 | -0.375 |  |
| Utilities | -0.426 | -0.519 | 0.145 |  |
| Physical activity | -0.238 | 0.178 |  | -0.724 |
| Sedentarism | -0.292 | -0.196 | 0.373 | 0.481 |
| Daily fruit & veg consumption | -0.144 | -0.407 | -0.186 | -0.476 |
| Salt consumption | -0.151 | 0.313 |  | 0.279 |
| Alcohol consumption | -0.225 | 0.324 | 0.496 | -0.171 |

The first principal component was negatively correlated with educational attainment (both for the interviewee and head of the household) and income, followed by working hours, while the second component presented higher loadings for density of inhabitants in the household and interviewee’s age (Table 2). Variables related to the usual lifestyle recommendations (level of physical activity, diet and alcohol consumption) did not have high loadings for the first two components, although they were the most correlated with the fourth component. Therefore, the first two components were mainly determined by socioeconomic variables while lifestyle influenced the third and fourth ones. The fifth and seventh component were mostly associated with salt and alcohol consumption, respectively, with the rest of the variables showing lower loadings (Additional File 3).

Superimposing smooth density estimates for Normal, Outlier or Extreme groups for fasting blood glucose residuals over the PCA biplot allowed us to explore how they differ in their characteristics for the explanatory variables. Figure 2 (a) shows distinct groups partially separated by the first and second Principal Components; there was a higher density of individuals with extreme residual values in the sector corresponding to a low income and low educational attainment (both for the interviewee and the head of the household). The higher density for the normal group was found in the region of higher educational attainment, higher income and lower age. No clear pattern was found regarding lifestyle variables, and Figure 2 (b) shows that the three groups mostly overlapped for the third and fourth Principal Components. No clear patterns were found for the rest of the components (Additional File 4). Similar results were obtained by using residuals of the linear regression between glycemia and age (not shown).

**Fig. 2.**
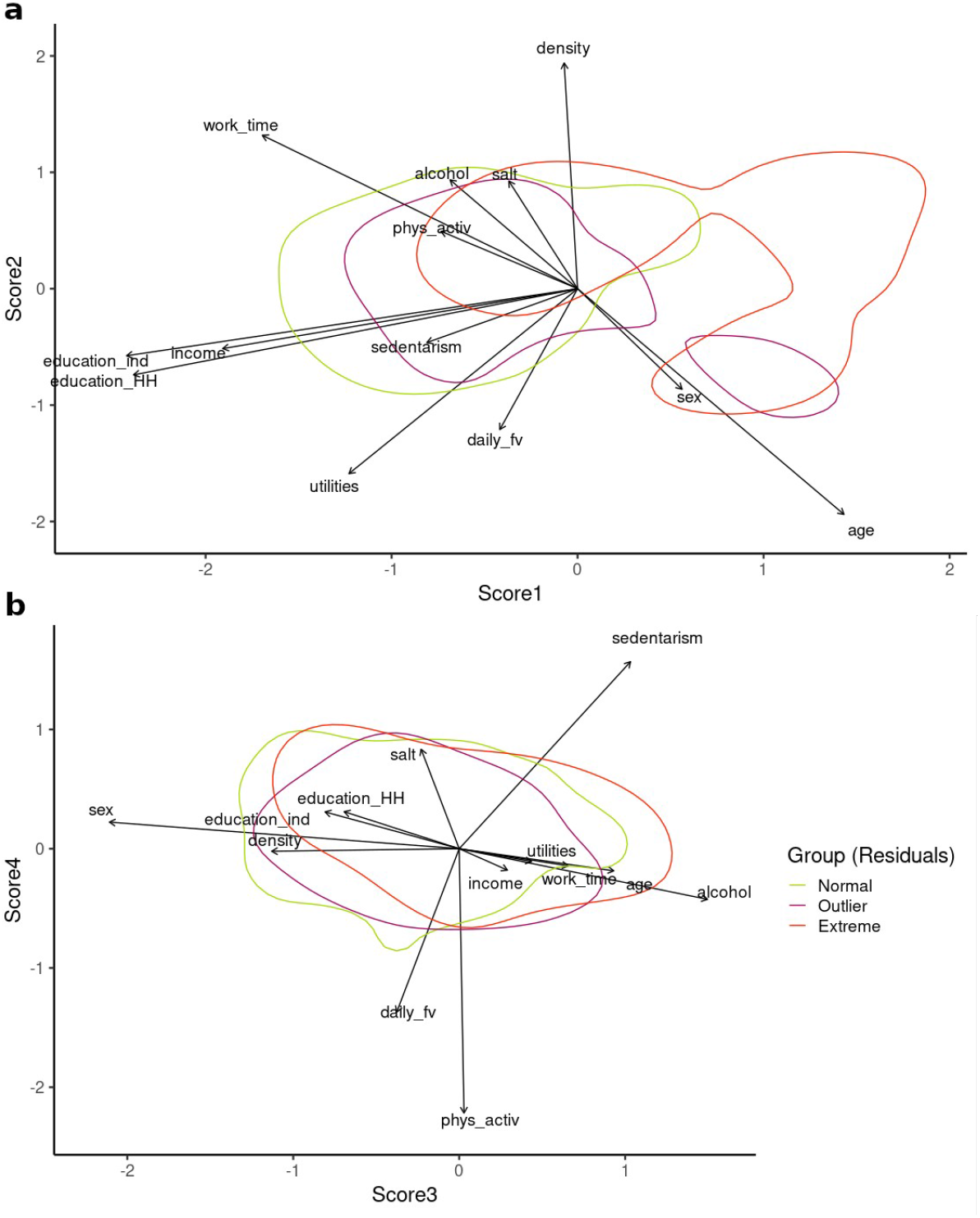
Curves for Normal, Outlier and Extreme glycemia residuals Biplots for the first and second components (a) and third and fourth components (b) of the PCA. Density curves for Normal, Outlier and Extreme groups (bins=2) are superimposed.

To analytically determine whether there were distinct regions of higher density of normal, outlier or extreme values for fasting blood glucose in the space determined by the Principal Components, we performed a PERMANOVA, after confirming no significant departures from homogeneity of multivariate dispersion in this multivariate space (F_2,4112_ = 0.729; p = 0.47). The PERMANOVA indicated that there were significant differences between Normal, Outlier and Extreme groups of blood glucose residuals for their position in the multidimensional space regarding all components retained in the PCA (F_2,4112_ = 20.95; p = 0.001). Indeed, the three groups differed significantly in pairwise comparisons (Table 3).

**Table 3.**
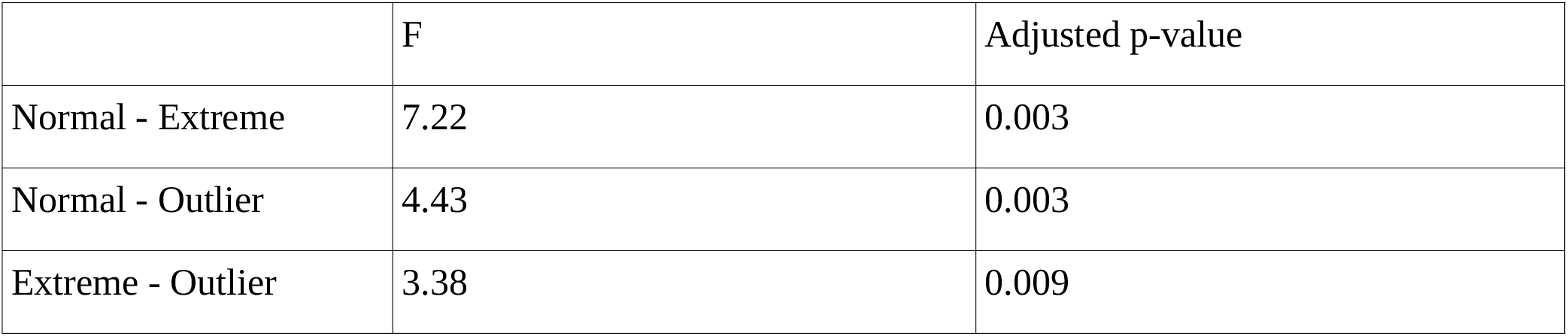
Results of the pairwise comparisons for the PERMANOVA test

The calculations of the running standard deviations showed a clear correlation between the first principal component and heterogeneity for fasting blood glucose (Figure 3), with the regions corresponding with lower income and educational attainment presenting neighborhoods with a higher variance for the trait. For the third and fourth components, a small region of high variance was found corresponding to the higher scores for the third component (higher alcohol consumption, older age, male gender), but no clear tendency was apparent overall (Figure 4). No defined pattern was observed either for the rest of the components (Additional File 5). These results did not change when correcting by running average (not shown).

**Fig. 3.**
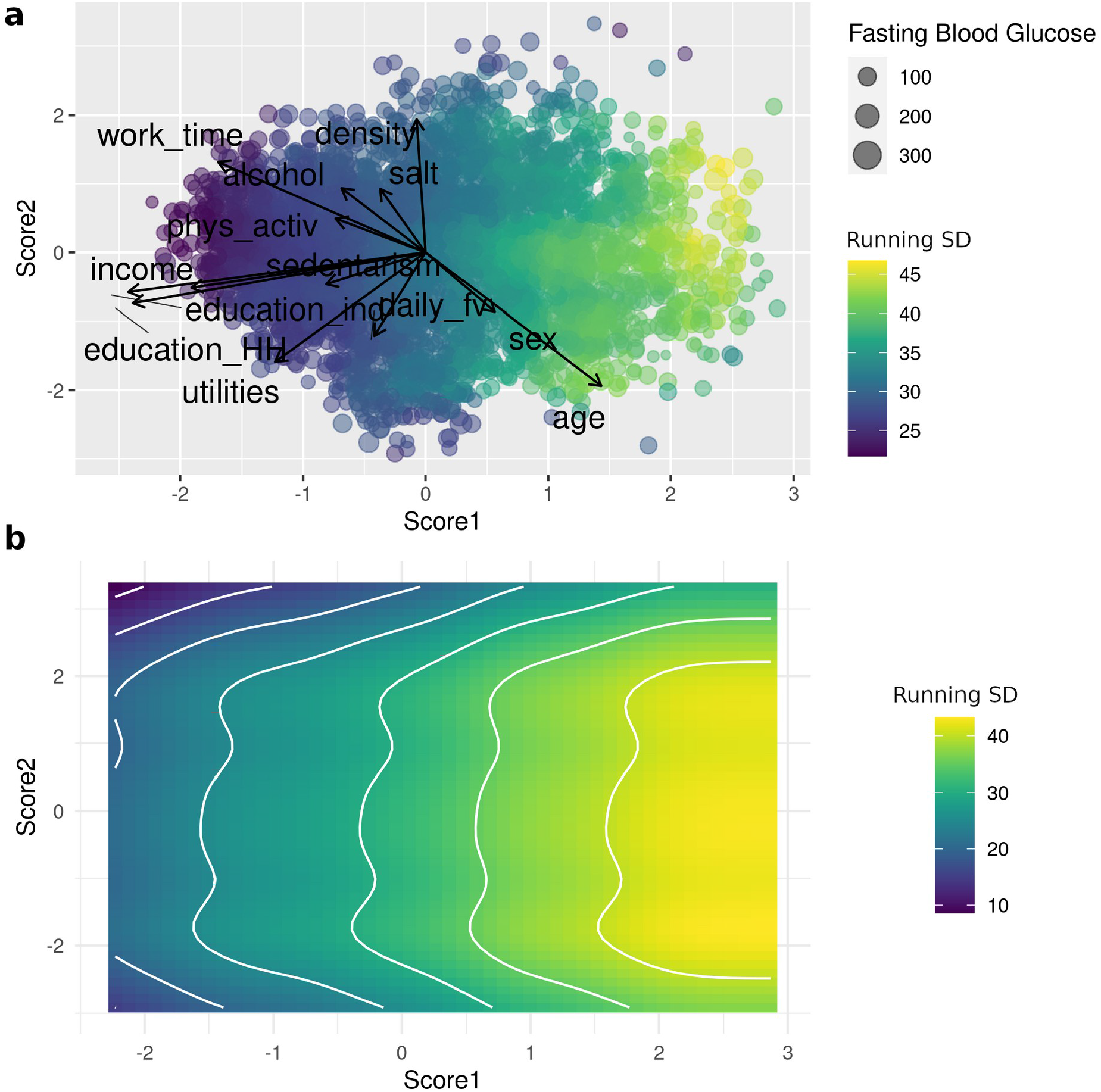
Running standard deviation for fasting glycemia in the first and second components of the PCA (a) Biplot for the first and second components of the PCA; point size correlates with fasting blood glucose levels (mg/dL) and color scale indicates levels of running standard deviation (mg/dL). (b) GAM smoothed curves for the running standard deviation.

**Fig. 4.**
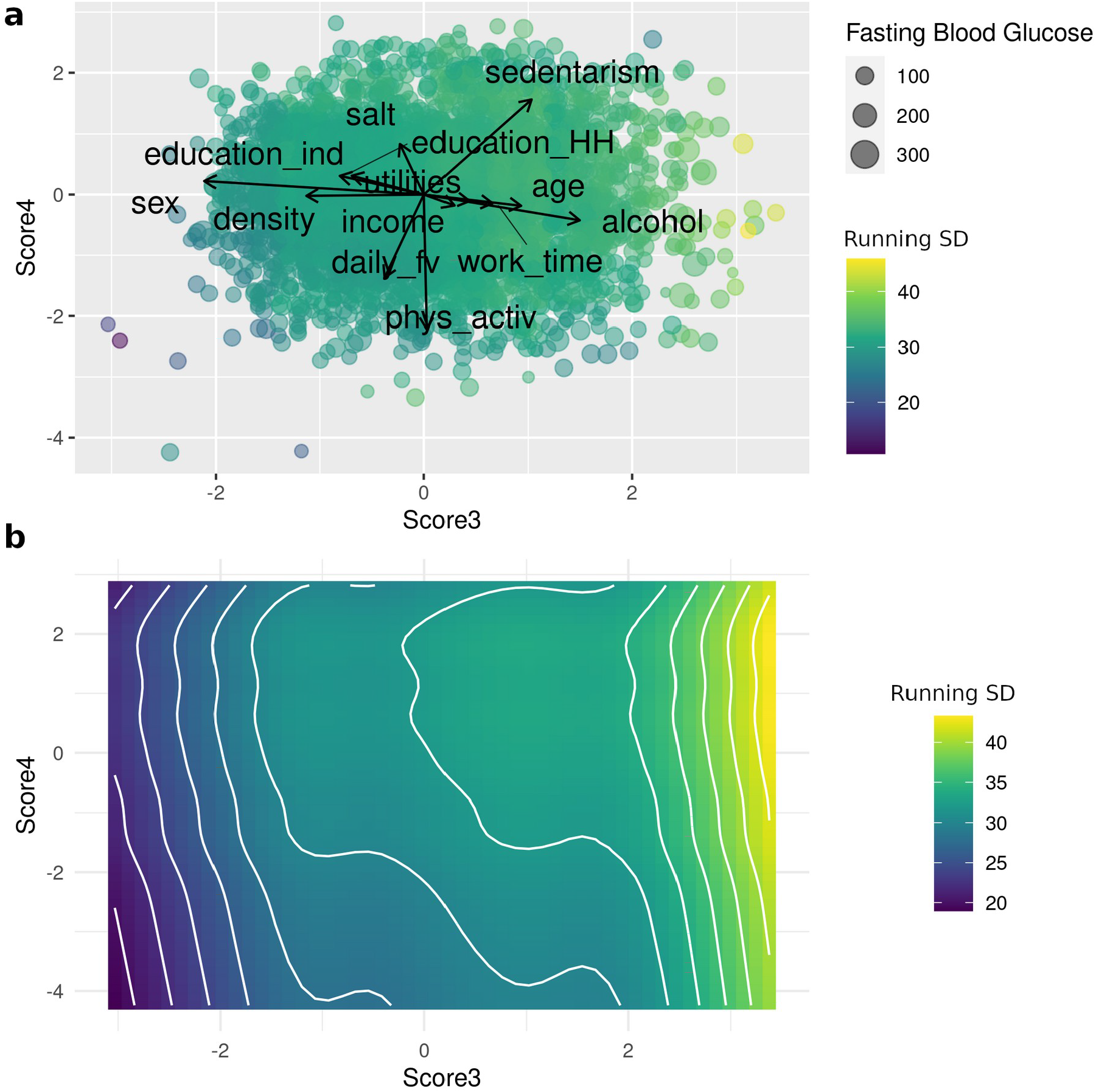
Running standard deviation for fasting glycemia in the third and fourth components of the PCA (a) Biplot for the third and fourth components of the PCA; point size correlates with fasting blood glucose levels (mg/dL) and color scale indicates levels of running standard deviation (mg/dL). (b) GAM smoothed curves for the running standard deviation.

As income and educational attainment appear as highly correlated relevant explanatory variables, we formed four income categories (see Materials and Methods) and estimated Coefficients of Variation of blood glycemia for all income and educational level combinations. Figure 5 shows the expected results: a trend for higher variability in low income groups. Also a reverse relationship between variability of glycemia and educational attainment can be found, although the latter is not clear in the higher and lower income groups, probably due to the scarcity of interviewees in the highest income – lowest educational attainment and lower income – higher educational attainment groups.

**Fig. 5.**
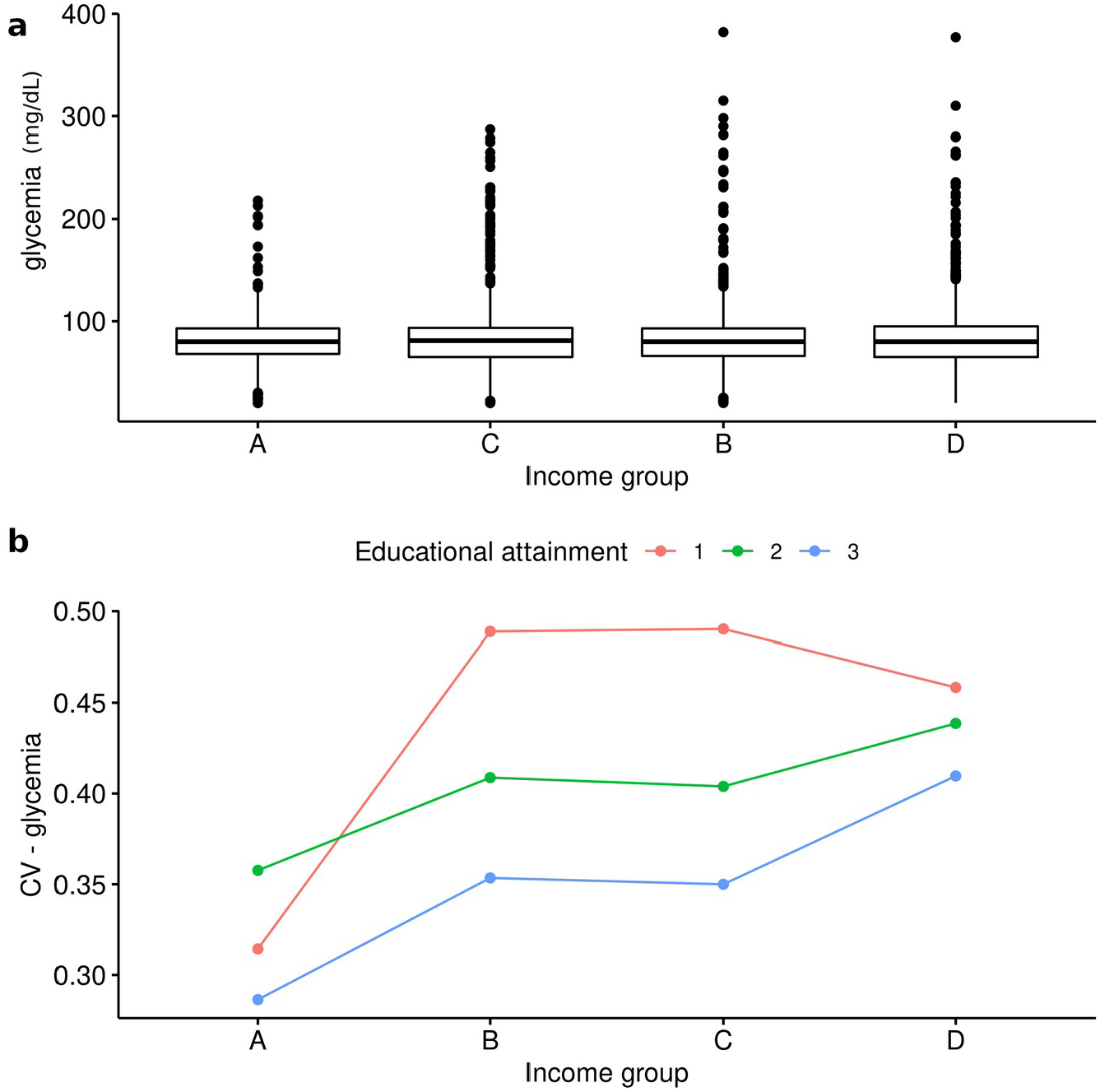
Fasting glycemia and Coefficients of Variation by income group Above: boxplots for values of glycemia (mg/dL) for the four income groups. Below: Coefficients of Variation for glycemia for the four income groups and the three levels of educational attainment.

Finally, to analytically test if the distribution within Normal, Outlier and Extreme groups for the residuals from the linear regression between glycemia and age is independent of these socioeconomic variables, we formed groups by combining categorized income with individual educational attainment and performed a G-test for independence. We found significant deviations from independence (G-test_df=22_ = 68.69; p = 1.06e-06), indicating that the proportion of individuals within the Normal, Outlier or Extreme groups depends on the income-educational attainment combination. The relative contributions to the statistic for each category can be found in Figure 6. For the high income and high education group, individuals in the Normal category for the regression between glycemia and age were more frequent than expected, while there was an excess of individuals at the Extreme and Outlier categories in the lower income and lower education groups. Interestingly, this trend was reversed in the low income and high education (C-3) group. However, we could not evaluate if this effect held for the higher and lower income groups, where lower and higher educational attainment, respectively, were not frequent.

**Fig. 6.**
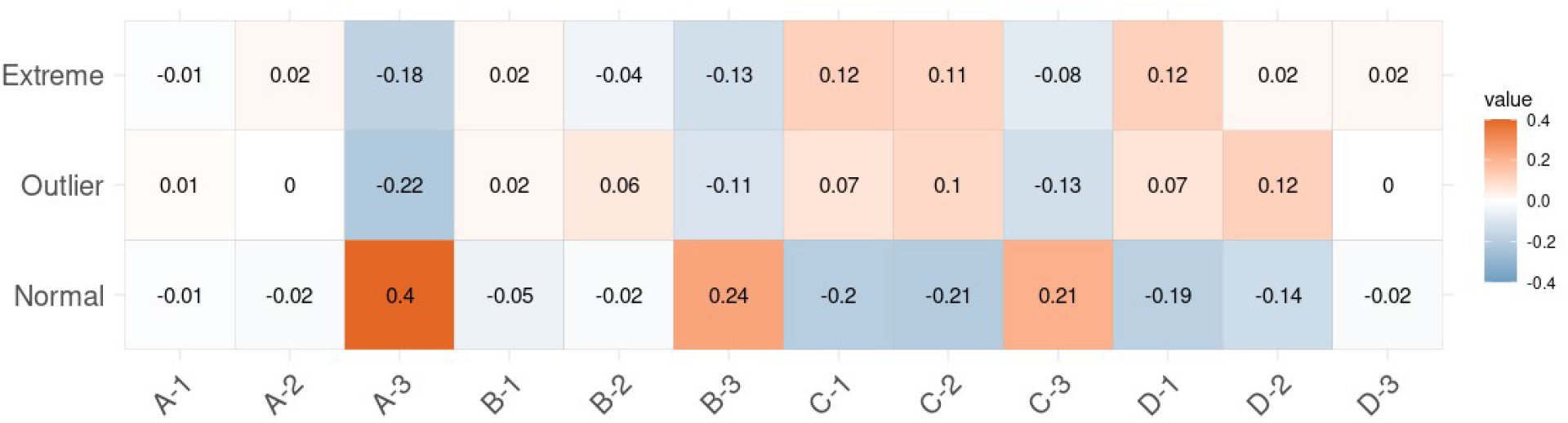
Results of the frequency analysis for categorized glycemia residuals Residuals relative to G-statistic value for all combinations of income and educational attainment. Red color indicates that the frequency of individuals in the group is higher than expected if blood glucose residual levels were independent of income and educational groups, while blue denotes a lower frequency than expected under the assumption of independence.

Finally, we wanted to analyze whether these statistical trends also had a clinical significance, meaning whether the distribution of blood glycemia values in the different income-education groups represented particular patterns of outcomes with impact on life quality. As expected, a higher proportion of individuals in the lower income and lower educational attainment groups presented levels of glycemia that can be categorized as prediabetes, diabetes or severe diabetes (Figure 7). It may be noted that hypoglycemia is also more prevalent in the lower income groups.

**Fig. 7.**
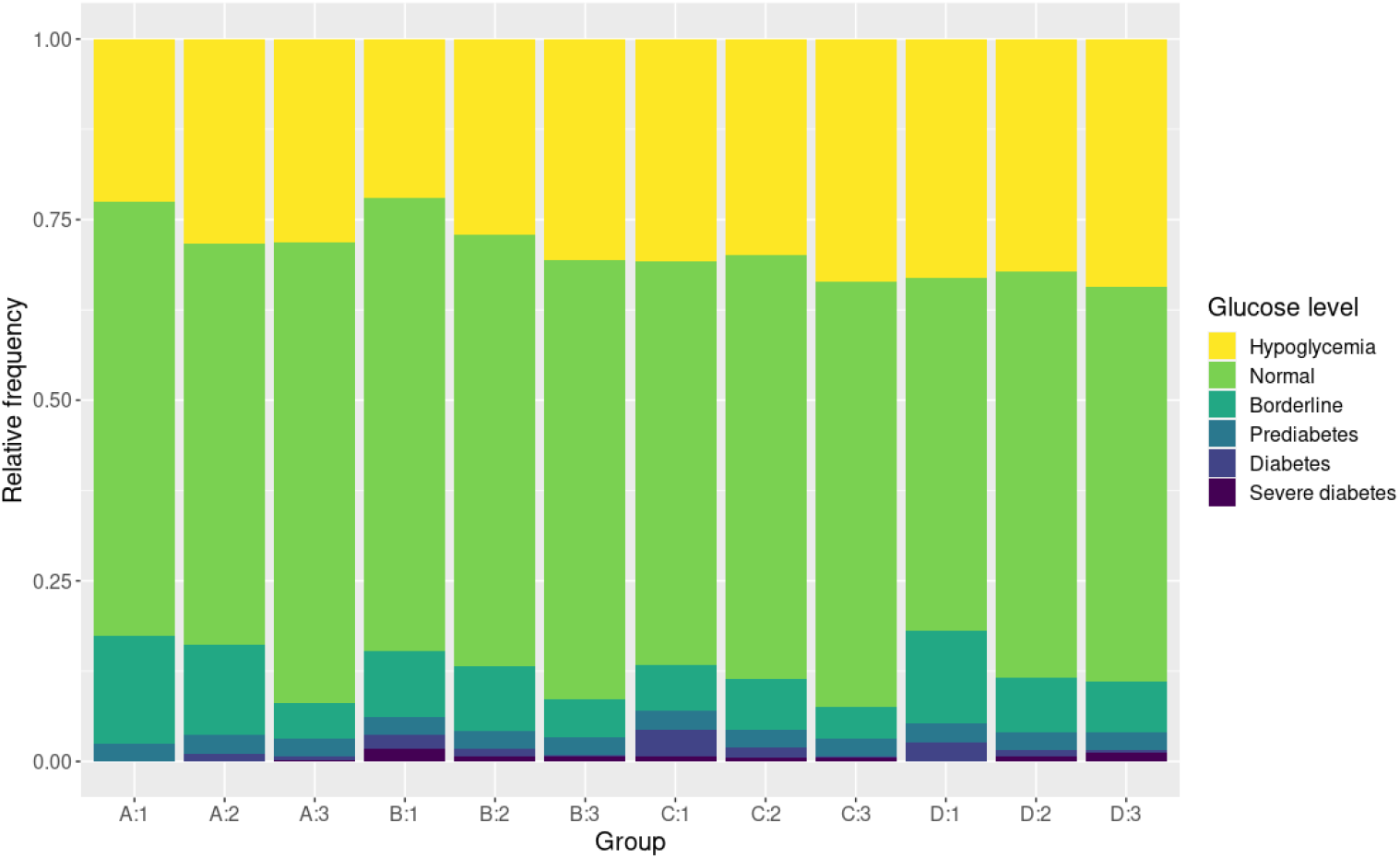
Levels of fasting glycemia per income and educational attainment Relative frequency of individuals presenting each glucose level at each income and educational attainment combination

## Discussion

Here we have shown how data from an Argentinian National Health Survey fit the hypothesis of phenotypic decanalization (in this case, of fasting blood glucose values) in response to socioeconomic variables, with a gradient in which the most disadvantaged groups show a greater variance than those in the best position as defined by income and educational attainment. Our results contrast with the dominant conception according to which socioeconomic situation acts through making the adoption of healthy habits not feasible [1, 3]. The data here analyzed show little to no evident relationship between lifestyle variables and variability for blood sugar values. It should be noted, however, that socioeconomic variables surely condition the adoption of healthy habits to some extent, as some answers to the survey indicate [19].

These results are relevant for several different reasons. First, they are important in the statistical sense because of the issues they raise as a corollary: predicting individual risk would be highly ineffective, as the augmentation of variance constitutes a group property. This is consistent with our inability to find any “traditional” model that would help explain or predict individual blood sugar values reliably. In fact, a higher group variance implies a heightened individual probability of both hyper and hypoglycemia. Therefore, if the decanalization hypothesis is true, a higher incidence of diabetes or high blood sugar could be found even without a change in means of glycemia by group, and this fact has important implications regarding statistical methods to search for factors affecting health and the design of population surveys.

But this finding is not only relevant in the statistical sense; these higher variances determine groups in which a higher proportion of individuals suffer from levels of blood glucose that are associated with pathology, or significant susceptibility to develop a health condition, as we have shown in Figure 7. These high fasting blood sugar values lower quality of life, increase the risk of short and long-term complications, some even life-threatening [1], and worsen the prognosis for several other conditions [4, 5, 38].

There is a number of advantages of this kind of analysis. First, it overcomes some of the most frequent critiques to traditional epidemiological research [39] such as that it does not propose mechanisms behind increased prevalence, and therefore the real causes of ill health may become invisibilized. Also, that it tends to disregard, minimize or simplify social and economic variables [40]. This is particularly troubling in the case of diseases affected by social or economic factors that may become masked by other variables more consistent with the idea of health as determined by individual (lifestyle or genetic) factors, therefore making the individuals responsible for their ill health, and promoting constant self-surveillance [41, 42]. In contrast, our approach includes a possible mechanism behind the increased prevalence of complex disease in vulnerable (or vulnerated [43]) populations that can accommodate a more holistic understanding of health and pathology processes, including the shifting social and economic relationships in the explanation of phenotypic changes [40], and also would lead to concrete strategies and public policy recommendations, without focusing on individual behaviors.

It should be noted that this approach is also not contingent on guidelines and rigid categorizations on healthy and ill individuals, which may be debatable, given the many interests and perspectives behind these discussions and the notions of health and disease themselves, and therefore result in heterogeneously adopted or changing criteria. In this sense, that this hypothesis does not rely in fixed boundaries between health and disease can be seen as an advantage and an opportunity to leave open the discussion on such boundaries.

in this sense, blood glucose determinations are an inestimable tool, as previous surveys only accounted for high blood glucose or diabetes by self-report as a categorical variable. Also, as they were performed in a probabilistic fashion, they provide a sample that is representative of the adult urban population of Argentina. Should this methodology be retained for future surveys, it will also allow for a follow-up of patterns of diabetes in the Argentinian population.

However, our analysis has some disadvantages and limitations. First of all, biochemical determinations were only carried out in interviewees from districts with a population higher than 150.000 [19]. Therefore, our conclusions can only apply to an urban population, as previously mentioned. Another limitation on the survey concerns age, since all interviewees were adults (age >= 18 years), although this could in fact be advantageous because it weakens the weight of type 1 diabetes as a confounding factor, as it is usually detected at an earlier age (see below).

Also, the survey lacked detailed information on the geographical location of interviewees. Even as we could detect groups defined by their multivariate socioeconomic space, geographical information would have allowed us to find “natural” groups, clusters of interviewees that, we would expect, shared a more similar environment. This data would probably condense the information on general quality of life that is not available in the survey in its present form (type of neighborhood, exposure to pollutants, or other stressful environmental conditions not covered by the questionnaire).

Other modifications that could be suggested to better characterize the individuals’ situation include gathering more detailed information on housing conditions, employment stability, unpaid workload and also subjective assessments on life quality.

Another downside of this approach is the inability to distinguish between type 1, type 2, and gestational diabetes. However, as mentioned above, type 1 diabetes is usually detected in childhood or adolescence [44] and has a low prevalence, with an estimate for Argentinian children and adolescents of 8.6 per 100,000 people [45] and of 15 per 100,000 people worldwide [46], and is also more likely to depend on genetic factors [47]. Therefore it is safe to assume that it should not affect our results to a great extent. On the other hand, since the Risk Factor Survey does not account for a possible pregnancy, a high blood sugar level caused by gestational diabetes cannot be ruled out. According to the survey, from the total of women with a self-reported history of diabetes, 21.6% were pregnant when diagnosed. However, it would probably not change our present results, as a low percentage of women would have been pregnant at the time of the survey, when measurements were taken. In fact, it has been estimated that type 2 diabetes represents up to 95% of the cases in developing countries [6], and therefore it is expected to account for the majority of the cases recorded in the survey.

Finally, our design does not allow us to determine causality, although the similar results seen by other researchers in different countries are suggestive that the effect of these particular factors in the distribution of health outcomes can be pervasive in different groups [14, 48]. Here, in the case of educational attainment and its apparent interaction with family income, we cannot differentiate between two possibilities: i) that lower educational attainment is indicative of a situation of long-term vulneration so that the relatively higher prevalence we have seen in middle-income individuals with lower educational attainment compared to those with higher education relates to this long-term effects; and/or ii) that higher educational attainment acts as a “protective” factor (for example, through increased social bonding, favoring autonomy and empowerment, and/or making it feasible to secure a steady job and therefore more stable living conditions). Indeed, other researchers have already proposed that access to higher education may counterbalance to some extent the heightened susceptibility to ill health due to low income ([14] and references). Our results are consistent with both explanations, although the pattern is less clear in the lower-income group, possibly because of the low number of individuals in the high educational attainment category, or else because even higher education cannot offset the stresses related to a family income below indigence threshold.

It should be noted, also, that even if income, and to some extent educational attainment, are recognized widely as affecting health in different contexts, our approach does not search for universal laws governing health. However, we argue that this methodology can be useful to detect local patterns of health outcomes, and that these insights should be integrated into a pluralistic framework including research at different levels, combining these more general quantitative analyses with in-depth, qualitative studies, to better understand the processes behind health disparities and how these are experienced by individuals and communities in different contexts [49].

To our knowledge, this is the first work that addresses a possible relationship between these socioeconomic variables and the decanalization hypothesis for fasting blood glucose. It is worth noting that the conclusions reached regarding public policy are very similar to the ones proposed by Marmot [14]: adequate living standards in the broad sense should be granted, which means reinforcing support to vulnerated groups. If the stresses related to lack of stability (of housing, working conditions, etc.) and a subordinate social position are responsible for the heightened incidence of diabetes, policies that focus on healthy habits and/or access to health care will not suffice to prevent further cases. Himmelstein et al. [13], who found results consistent with the decanalization hypothesis for blood pressure, suggest reducing the exposure to these stressors and protecting at-risk populations from environmental fluctuations. According to them, the strengthening of social networks and the access to behavioral “buffering” pathways would partly unload the burden posed on the physiological subsystem. Marmot [14] similarly argues that empowerment acts as a protecting factor and suggests policies to improve income, education, housing stability, working conditions, etc. Our results support these suggestions. Although these policies would imply an increase in spending not directly focused on health care, such measures may have more meaningful impacts than current health policies for diseases like type 2 diabetes, which constitute a growing economic burden both for individuals and health systems [50].

We believe the decanalization hypothesis is a promising model for the study of the complex and historically changing patterns of susceptibility to ill health, particularly to chronic multi-factorial ailments related to homeostasis, which can incorporate biological (ecological, developmental, evolutionary) and socioeconomic determinants. We believe our approach, however, can be improved. Changes in data gathering methodology may benefit from interdisciplinary work to incorporate useful variables to further test this hypothesis, as current survey designs and statistical methods are not well-tailored to this end. However, this is not the only reason why we want to highlight the importance of the dialogue between social, biological, and health sciences. The limitations generated by disciplinary fragmentation on epidemiology have already been described, and different counter-strategies proposed [12, 39, 40, 49]. The notion of human populations as immersed in their socio-ecosystems, constrained by social and material conditions that are affected by a myriad of factors with a particular history and contexts, is necessary to understand the processes behind health and disease through the incorporation of different approaches and methodologies. This requires avoiding the social-biological dichotomy and overcoming disciplinary fragmentation. This issue has been raised for several subdisciplines of biology [51] including those involved in human health [12, 40, 52, 53]. But moreover, our own notions and concepts of health and disease and the frameworks used to understand the complexity of the processes behind them also require problematizing and rethinking, and interdisciplinary work may fuel constant revisions of our approaches and substantiate a more pluralistic take on these issues [49].

## Data Availability

All data produced in the present work are contained in the manuscript and additional files

https://www.indec.gob.ar/indec/web/Institucional-Indec-BasesDeDatos-2

## Statements

All authors prepared, read and agreed with the content, gave consent to submit and obtained consent from the responsible authorities at their respective institutes.

The authors declare they have no actual or potential competing financial interests. No specific funding was destined to the preparation of this manuscript and the authors have no affiliations with or involvement in any organization or entity with any financial interest or non-financial interest in the subject matter or materials discussed in this manuscript.

This research involves only the use of public anonymized data and therefore was granted exemption from ethics committee approval. The study complies with the principles of the Declaration of Helsinki.

## Additional file captions

Additional File 1. Data from the National Health Survey for all individuals retained.

Additional File 2. Summary of the regressions performed to model blood glycemia data. Method, proportion of variance explained and the detailed models (in R language) are provided

Additional File 3. Loadings for all the variables in the 8 principal components generated

Additional File 4. Curves for Normal, Outlier and Extreme glycemia residuals. Biplots for the components 5, 6, 7 and 8 of the PCA. Density curves for Normal, Outlier and Extreme groups (bins=2) are superimposed.

Additional File 5. Running standard deviation for fasting glycemia. Biplots for the components 5, 6, 7 and 8 of the PCA; point size correlates with fasting blood glucose levels (mg/dL) and color scale indicates levels of running standard deviation (mg/dL). GAM smoothed curves for the running standard deviation are also provided.

