## Additional File 3 for "Testing the phenotypic decanalization hypothesis: social determinants of hyperglycemia and type 2 diabetes in adult urban Argentinian population"

Additional File 3. Loadings for all the variables in the 8 principal components generated

```
Loadings (cutoff = 0.1):
              Comp1  Comp2  Comp3  Comp4  Comp5  Comp6  Comp7  Comp8
education_ind -0.808 -0.192 -0.269  0.103         0.256  0.155
education_HH  -0.796 -0.247 -0.231  0.104         0.248  0.161
income        -0.636 -0.171                      -0.273 -0.265  0.352
work_time     -0.565  0.439  0.219         0.242
density               0.647 -0.377         0.267 -0.234 -0.134  0.283
utilities     -0.410 -0.530  0.145        -0.151        -0.417  0.220
age            0.477 -0.646  0.310
sex            0.187 -0.288 -0.703               -0.108  0.192
phys_activ    -0.245  0.164        -0.739 -0.140        -0.146 -0.441
sedentarism   -0.270 -0.154  0.344  0.523  0.312 -0.349  0.130 -0.393
salt          -0.123  0.309         0.278 -0.753 -0.421 -0.114 -0.147
daily_fv      -0.139 -0.403 -0.126 -0.457  0.152 -0.578  0.333
alcohol       -0.229  0.312  0.499 -0.142 -0.272         0.549  0.361

Importance (Variance Accounted For):
                 Comp1   Comp2   Comp3   Comp4   Comp5   Comp6  Comp7   Comp8
Eigenvalues     2.6622  1.9251  1.3254  1.1636  0.9613  0.9068  0.822  0.7731
VAF            20.4782 14.8087 10.1957  8.9510  7.3945  6.9751  6.323  5.9470
Cumulative VAF 20.4800 35.2900 45.4800 54.4300 61.8300 68.8000 75.130 81.0700
```
