## Supplementary figures and images for "Testing the phenotypic decanalization hypothesis: social determinants of hyperglycemia and type 2 diabetes in adult urban Argentinian population"

### Additional File 4

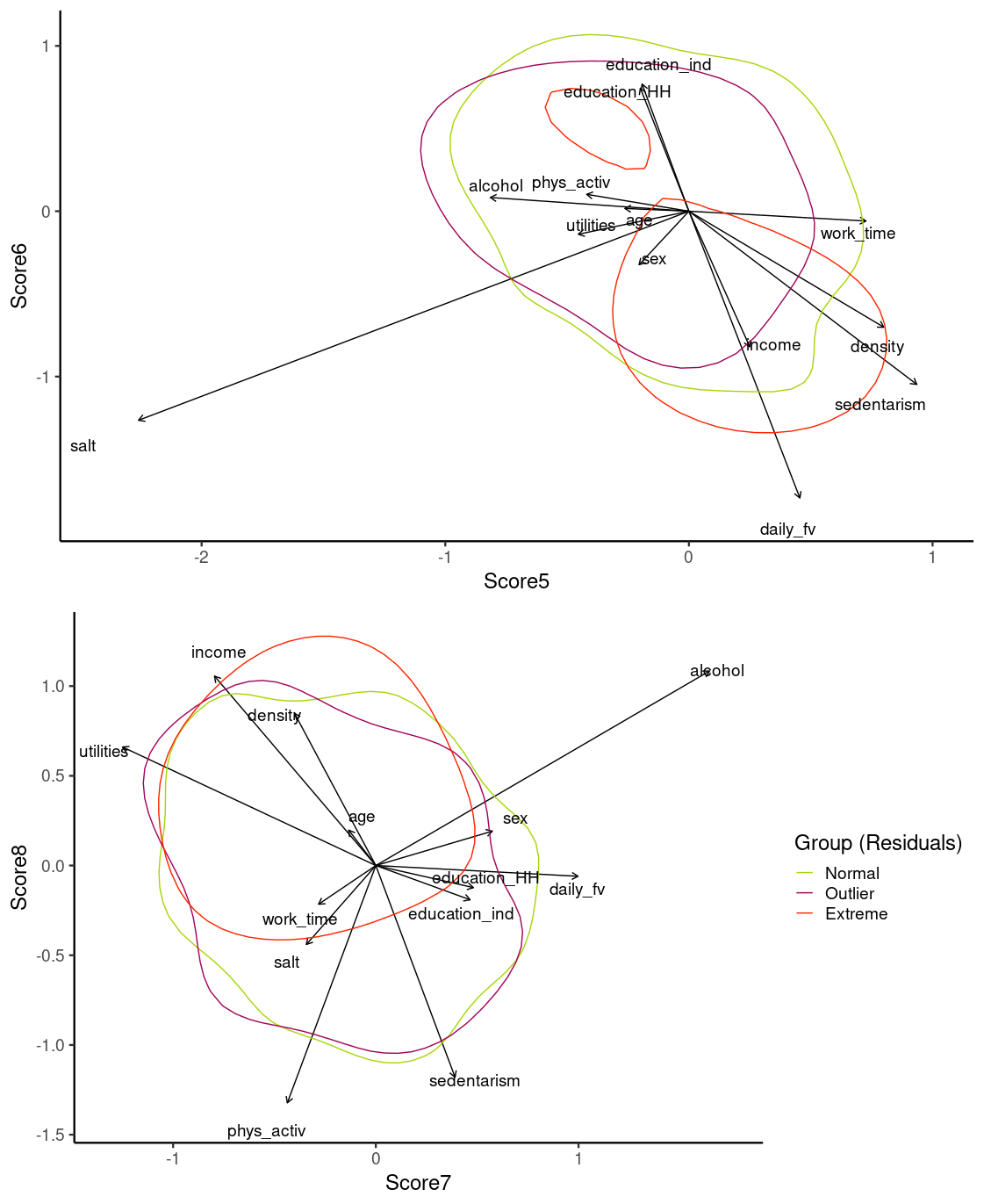

### Additional File 5

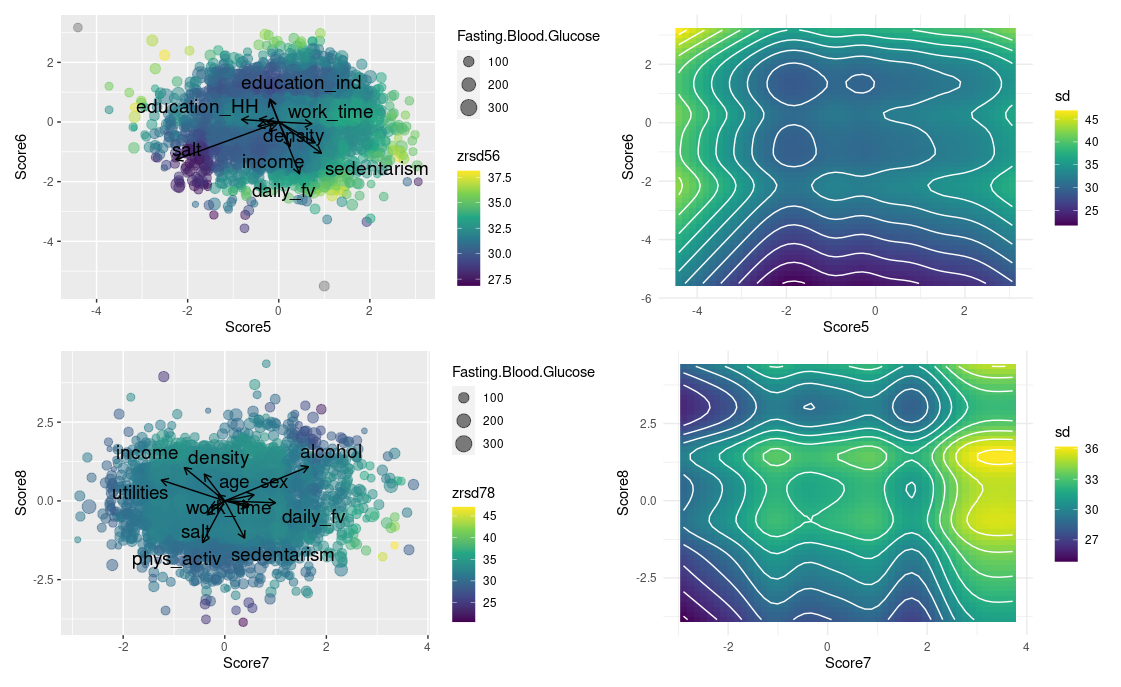
